# Associations between SARS-CoV-2 Infection and Multidimensional Sleep Health

**DOI:** 10.64898/2026.02.19.26346546

**Authors:** Salma Batool-Anwar, Matthew D. Weaver, Mark É. Czeisler, Lauren A. Booker, Mark E. Howard, Melinda L. Jackson, Christine F. McDonald, Rebecca Robbins, Prerna Varma, Shantha M.W. Rajaratnam, Charles A. Czeisler, Stuart F. Quan

## Abstract

**Puhrpose:** To evaluate the short- and long-term cross-sectional associations between COVID-19 infection and multidimensional sleep health.

**Methods:** Data from the COVID-19 Outbreak Public Evaluation (COPE) initiative were used to examine the association between a novel multidimensional sleep health measure (COPE Multidimensional Sleep Health Scale, CMSHS) modeled from the RuSATED instrument and (1) COVID-19 infection and (2) post-acute sequelae of SARS-CoV-2 infection (PASC).

**Results:** Data from 11,326 respondents were used for this study. The cohort was comprised of 51% women, 61% non-Hispanic White, and 17% Hispanic adults. COVID-19 infection was more prevalent among participants who had not received a booster vaccination (55.4% vs. 30.2%, p<0.001); the number of comorbid conditions was higher among those who had been infected (2.2% vs. 1.7%, p<0.001). Participants with COVID-19 infection had significantly lower CMSHS scores indicative of worse sleep health compared with uninfected participants (3.52 ± 1.37 vs. 3.78 ± 1.30; p < 0.001). Participants with PASC had lower CMSHS scores in comparison to those without PASC (2.72 ± 1.30 vs. 3.82 ± 1.28, p<0.001). In adjusted models, a progressive decline in CMSHS scores was observed over 12 months following infection (3.52 ± 0.05 vs. 2.98 ± 0.04; p < 0.001 for <1 month vs. 6–12 months).

**Conclusion:** Compared with uninfected individuals, multidimensional sleep health was worse among persons who had a COVID-19 infection. Individuals with PASC had greater and persistent reductions in sleep health for up to 12 months post-infection.

**Brief summary:**

1. Several studies have examined the negative effects of COVID-19 on sleep, however the effects of COVID-19 infection on multidimensional sleep health remain poorly understood as do these associations over time. Using a large, population-based cohort, this study evaluates short- and long-term effects of Covid-19 infection on overall sleep health.
2. The study provides evidence that COVID-19 infection is associated with impairments in overall sleep health, with effects persisting up to 12 months post-infection. The findings in this study demonstrate that poor sleep health is an important long-term consequence of COVID-19 infection and emphasizes the need for sleep assessment among patients affected by COVID-19.

## Introduction

Since the beginning of the COVID-19 pandemic, hundreds of millions of people globally have been infected by SARS-CoV-2 [1]. A significant proportion of them experienced a variety of symptoms persisting beyond the initial infection, recognized as Post-Acute Sequelae of SARS-CoV-2 (PASC, long COVID). It has been estimated that approximately 100 million people suffer from PASC and that the total economic burden attributable to PASC is estimated to be approximately 2.6-3.7 trillion dollars [2]. Besides fatigue, generalized weakness, and anxiety, poor sleep is frequently experienced among people with PASC [3],[4]. In addition to the direct effects of infection, isolation and quarantine may exacerbate sleep difficulties [5, 6]. Moreover, an increase in symptoms among those with preexisting sleep disorders was also seen during the COVID-19 pandemic [7]. However, as documented in a systematic review of 250 studies worldwide, most reports of sleep disturbances in PASC are either non-specific, cross-sectional, or focused on a single sleep condition such as insomnia [8]; reports of the evolution of sleep disturbances over an extended time course after acute COVID-19 infection are less common [4].

Over the past decade, sleep researchers have increasingly conceptualized sleep health as containing multiple dimensions, recognizing that simultaneous consideration of multiple aspects of sleep is more informative than isolated assessments of individual dimensions [9]. As proposed by Buysse [9], overall sleep health can be determined by evaluating the following six dimensions: sleep regularity, subjective satisfaction, appropriate timing, adequate duration, high sleep efficiency, and sustained alertness during the day. To operationalize its assessment, the RuSATED instrument was developed to quantify overall sleep health and its six dimensions [9, 10]. The instrument has been shown to discriminate between good and poor sleep health in comparative studies of populations with and without depression [11], hypertension [12], and cardiovascular disease [12],[13]. Using the SATED and Ru-SATED questionnaires, in a recent repeated cross-sectional study examining sleep health trends following COVID-19 pandemic in a general population, the authors demonstrated uneven sleep health improvement across several sleep dimensions [14].

Despite evidence suggesting an association between COVID-19 infection and poor sleep, little is known about multidimensional sleep health among persons with previous COVID-19 infection as well as those who subsequently developed PASC. We also know little about these associations over time. Using the data from COVID-19 Outbreak Public Evaluation (COPE) initiative, we evaluated the effect of COVID-19 infection and PASC on sleep health and its component dimensions. We hypothesized that multidimensional sleep health as represented by the six concepts comprising the RuSATED scale would be lower among people with previous COVID-19 infection and in those with PASC than among individuals without prior SARS-CoV-2 infection, and that poor sleep health in those with PASC would persist over a prolonged period of time.

## Methods

### Study Design and Participants

From March 10 to October 15, 2022, the COVID-19 Outbreak Public Evaluation (COPE) Initiative (https://www.thecopeinitiative.org) administered five waves of cross-sectional surveys consisting of approximately 5,000 participants focused on accumulating data on the prevalence and sequelae of COVID-19 infection in the United States (U.S.). Dates of administration were Wave 1 (March 10-30, 2022), Wave 2 (April 4-May 1, 2022), Wave 3 (May 4-June 2, 2022), Wave 4 (August 1-18, 2022), Wave 5 (September 26-October 15, 2022). Recruitment strategy has been described previously [15]; it attempted to create samples that approximated population estimates for age, sex, race, and ethnicity based on the 2020 U.S. census. Questions used to create the sleep health assessment in this study were present on the 1^st^, 3^rd^, and 5^th^ waves and form the basis of our analysis. Surveys were conducted online by Qualtrics, LLC (Provo, Utah, and Seattle, Washington, U.S.). Informed consent was obtained electronically. The study was approved by the Monash University Human Research Ethics Committee (Study #24036).

### Survey Items

Participants self-reported their demographic, anthropometric, and socio-economic information, including age, race, ethnicity, sex, height and weight, education level, employment status, and household income. They were classified as having had COVID-19 if they endorsed having been tested positive for COVID-19 or were confident that they had been infected. They also provided details on the number of COVID-19 infections they had experienced (0 up to 4) and the number of COVID-19 vaccinations received. Information regarding then-current and past medical conditions was obtained by answers to the following question: “Have you ever been diagnosed with any of the following conditions? insomnia, high blood pressure, cardiovascular disease, gastrointestinal disorder, cancer, chronic kidney disease, liver disease, sickle cell disease, chronic obstructive pulmonary disease, and asthma.” Response options allowed for affirming the condition and whether participants were treated currently or in the past. Additionally, as done in our previous studies [15–17], the presence of obstructive sleep apnea was determined by asking participants if they had either of the following combination of symptoms: 1) snoring “three or more times a week” and witnessed apnea or sleepiness “once or twice a week”; 2) witnessed apnea and sleepiness “once or twice a week”.

The following survey items were used to derive an adaptation of the RuSATED multidimensional sleep health scale [9, 10]: Sleep regularity was assessed using responses from the following two questions: “During the past month, how has the COVID-19 pandemic affected the following, if at all? Sleeping at regular hours and Time in Bed”. Response options to both questions were “More than usual”, “Less than usual”, “Same as usual”. Sleep satisfaction was interpreted to be equivalent to sleep quality and was ascertained from answers to the question: “During the past month, how would you rate your sleep quality overall”? Response options were “Very good”, “Fairly good”, “Fairly bad” or “Very bad”. Level of alertness was assessed by responses to the following question concerning sleepiness: “During the past month, how often have you had trouble staying awake while driving, eating meals, or engaging in social activity?” Response options were “Not during the past month”, “Less than once a week”, “Once or twice a week”, or “Three or more times a week”. Sleep timing was determined by responses to the following questions: “During the past month, when have you usually gone to bed at night?” and “During the past month, when have you usually gotten up in the morning?”. Sleep efficiency and duration were calculated by responses to questions regarding the number of hours spent trying to sleep and the number of hours actually slept.

For participants who reported having had a positive COVID-19 test or were confident of having a COVID-19 infection, a series of questions were asked to ascertain the presence and duration of each of 13 symptoms used to define PASC (*vide infra*). At the following time points after COVID-19 infection, <1 month, 1-3 months, 3-6 months, and 6-12 months, participants who had been classified as having had COVID-19 were asked to recall if they had experienced each of the symptoms.

### Data Analyses

#### Exposures

Classification as positive for COVID-19 infection was defined as a self-report of having tested positive or being confident of having been infected. For these analyses, only participants who had never been infected or those with a single infection were included because inclusion of those with multiple infections would have confounded the interpretation of the longitudinal associations between sleep health and PASC. Vaccination status was dichotomized as Boosted (>2 vaccinations) or Not Boosted (≤2 vaccinations). Comorbid medical conditions were defined as currently having the condition whether treated or untreated. As in our previous studies [15, 16, 18], the effect of comorbid medical conditions was evaluated by summing the number of conditions reported by the participant to create a comorbidity index (minimum value 0, maximum value 10). Body mass index (BMI) was calculated using self-reported height and weight as kg/m^2^. Socioeconomic covariates were dichotomized as follows: employment (retired vs. not retired), education (high school or less vs. some college) and income in U.S. Dollars (<$50,000 vs ≥$50,000).

The presence of PASC was determined using algorithms that we have previously published. Briefly, 2 definitions of PASC developed in prior analyses of the COPE cohort were used in this study [18]. The first was the presence for at least 3 months of at least 3 of 13 symptoms (Supplemental Table S1) that are often observed after a COVID-19 infection (COPE definition). The second was the presence of at least 1 of the aforementioned symptoms persisting for at least 3 months after infection as suggested by the United Kingdom’s National Institute for Health and Care Excellence (NICE definition).

#### COPE Multidimensional Sleep Health Scale (CMSHS)

The multidimensional sleep health score used in this study was designed based on tThe multidimensional RuSATED sleep health scale, which is composed of 6 sleep domains: Regularity, Satisfaction, Alertness, Timing, Efficiency, and Duration, each assessed with a 5-point Likert scale. RuSATED responses are summated to yield a maximum score of 30, with higher scores indicative of better overall sleep health [10]. Recently, a modification of RuSATED was developed using sleep diary and actigraphy derived data for which cutoff values were proposed to delineate “good” versus “poor” behavior for each dimension [12]. This yielded a 6 point scale ranging from 0 (very poor sleep health) to 6 (excellent sleep health). The modified scale was shown to associated with metrics of cardiometabolic activity as well as to vary appropriately across several other medical conditions [12]. For our study, we used a similar approach to assign component scores using cut-points as previously validated where applicable as follows:

Regularity: Regular (=1) for responses of “Same as usual” to both questions; Irregular (=0) for responses of either “More than usual” or “Less than usual” to at least one of the questions
Satisfaction: Satisfied (=1) for responses to the question regarding sleep quality of either “Very good” or “Fairly good”; Not Satisfied (=0) for responses of either “Fairly bad” or “Very bad”
Alertness: Alert (=1) for responses to the sleepiness question of either “Not during the past month” or “Less than once a week”; Not Alert (=0) for responses of “Once or twice a week”, or “Three or more times a week”
Timing: Good (=1) for sleep midpoint between 0224 and 0330 hours; Poor(=0) for all other midpoints.
Efficiency: Good (=1) for sleep efficiency >83%; Poor (=0) for sleep efficiency ≤ 83%.
Duration: Good (=1) for sleep duration between 5.33 and 7.10 hours; Poor (=0) for all other sleep durations

An overall multidimensional sleep health score (CMSHS) was calculated by summing the scores for each component (minimum=0, maximum=6 with higher scores indicative of better sleep health). To provide validation that the CMSHS could reflect levels of sleep health, we assessed whether the CMSHS score varied appropriately across medical conditions known to be associated with poor sleep health by comparing mean CMSHS scores between participants with and without the medical conditions comprising the comorbidity index. In addition, the areas under receiver operating curves (AUC) were calculated to assess the strength of the association between presence or absence of these conditions and CMSHS scores. To determine whether CMSHS was associated with COVID-19 infection, a comparison also was made between participants with and without a history of COVID-19 infection.

Missing data related to the components of CMSHS were present in as many as 18.9% of cases which would have resulted in exclusion of these cases in multivariate analyses. Missingness was assessed to be at random. Therefore, multiple imputation by chained equations was employed to generate replacement values. Comparison of the imputed dataset with the original dataset revealed no outliers in the imputed dataset, and the means of the same variables between the two datasets were comparable.

Using the imputed dataset, general linear models were used to determine whether CMSHS scores were associated with the COPE and NICE definitions of PASC. For each definition, a baseline model was constructed consisting only of the presence or absence of PASC. We then developed increasingly complex models by sequentially including demographics and anthropometric factors, comorbidities, boosted vaccination status, and socioeconomic factors.

Changes in CMSHS scores as a function of time after COVID-19 infection were evaluated using general linear models with the imputed dataset. Initially, a baseline model was constructed consisting only of a variable defining a No PASC control and timepoints after COVID-19 infection. Increasingly complex models with the addition of demographics and anthropometric factors, comorbidities, boosted vaccination status, and socioeconomic factors then were developed.

Summary data for continuous variables are reported as their respective means and standard deviations (SD) or standard errors (SE), and for categorical and ordinal variables as their percentages. For comparisons among multiple mean values, a Sidek correction was applied. A p<0.05 was considered statistically significant. All analyses were conducted using SPSS version 28 (IBM, Armonk, NY).

## Results

Table 1 contains the characteristics of the study population. Overall, there were 11,326 participants in Waves 1, 3, and 5 who provided data concerning the number of COVID-19 infections they had experienced. Of these participants, 4,328 had only one infection and 5,574 had never had an infection. In comparison to the cohort who had never been infected, those who had only one infection were younger (43.4 ± 16.9 vs. 48.7 ± 18.1 years, p<0.001) and more likely to be women (54.2 vs. 49.8%, p<0.001). There was also a greater proportion of Hispanic adults and a lower proportion of Asian adults among those with one infection. Participants with previous COVID-19 infection also had a greater number of comorbidities and a higher BMI. They were less likely to have received a COVID-19 booster, to be retired and to have an income below the poverty level. Educational attainment was not different between the COVID-19 infection and non-infected groups.

**Table 1:** Characteristics of the Study Population.

|  | Overall |  | No COVID-19 Infection |  | One COVID-19 Infection |  | p* |
| --- | --- | --- | --- | --- | --- | --- | --- |
|  | N=11326<sup>†</sup> |  | N=5574 |  | N=4328 |  |  |
|  | N | Mean ± SD / % | N | Mean ± SD / % | N | Mean ± SD / % |  |
| **Age** | 11326 | 45.6 ± 17.6 | 5574 | 48.7 ± 18.1 | 4328 | 43.4 ± 16.9 | <0.001 |
| **Sex** | 11255 |  | 5547 |  | 4295 |  |  |
| Men | 5503 | 48.9 | 2783 | 50.2 | 1968 | 45.8 | <0.001 |
| Women | 5752 | 51.1 | 2764 | 49.8 | 2327 | 54.2 |  |
| **Race/Ethnicity** | 11323 |  | 5571 |  | 4328 |  |  |
| Non-Hispanic White | 6945 | 61.3 | 3428 | 61.5 | 2669 | 61.7 | <0.001 |
| Black | 1129 | 10.0 | 620 | 11.1 | 386 | 8.9 |  |
| Asian | 633 | 5.6 | 371 | 6.7 | 210 | 4.9 |  |
| Hispanic | 2024 | 17.9 | 847 | 15.2 | 856 | 19.8 |  |
| Other Non-Hispanic | 592 | 5.2 | 305 | 5.5 | 207 | 4.8 |  |
| **Number of Comorbidities** | 11326 | 2.1 ± 2.8 | 5574 | 1.7 ± 2.3 | 4328 | 2.2 ± 3.0 | <0.001 |
| **Body Mass Index kg/m<sup>2</sup>** | 10362 | 27.7 ± 6.1 | 5189 | 27.6 ± 5.9 | 3949 | 27.9 ± 6.2 | 0.018 |
| **COVID Vaccine Booster** | 10678 |  | 5191 |  | 4114 |  |  |
| Yes | 3961 | 37.1 | 2317 | 55.4 | 1241 | 30.2 | <0.001 |
| No | 6717 | 62.9 | 2874 | 44.6 | 2873 | 69.8 |  |
| **Employment** | 10749 |  |  |  |  |  |  |
| Full/Part-Time | 6427 | 59.8 | 2802 | 53.0 | 2652 | 64.4 | <0.001 |
| Retired/Other | 4322 | 40.2 | 2481 | 47.0 | 1466 | 35.6 |  |
| **Income** | 10922 |  | 5340 |  | 4197 |  |  |
| <\$50,000 | 4639 | 42.5 | 2378 | 44.5 | 1759 | 41.9 | 0.010 |
| ≥\$50,000 | 6283 | 57.5 | 2962 | 55.5 | 2438 | 58.1 | |
| **Education** | 11326 |  | 5574 |  | 4328 |  |  |
| ≤High School | 2759 | 24.4 | 1343 | 24.1 | 1066 | 24.6 | 0.537 |
| ≥Some College | 8567 | 75.6 | 4231 | 75.9 | 3262 | 75.4 |  |

\*Comparison of No COVID-19 to One COVID-19 Infection
<sup>†</sup>Includes participants with >1 COVID-19 Infection

Shown in Table 2 are the CMSHS scores for a spectrum of chronic medical conditions. For all of these conditions, the CMSHS scores were higher, indicating better sleep health, in those without the condition in comparison to those with the condition. Furthermore, the AUC values for asthma, chronic obstructive lung disease, sickle cell disease, insomnia, and hypertension were greater than 0.7.

**Table 2:**
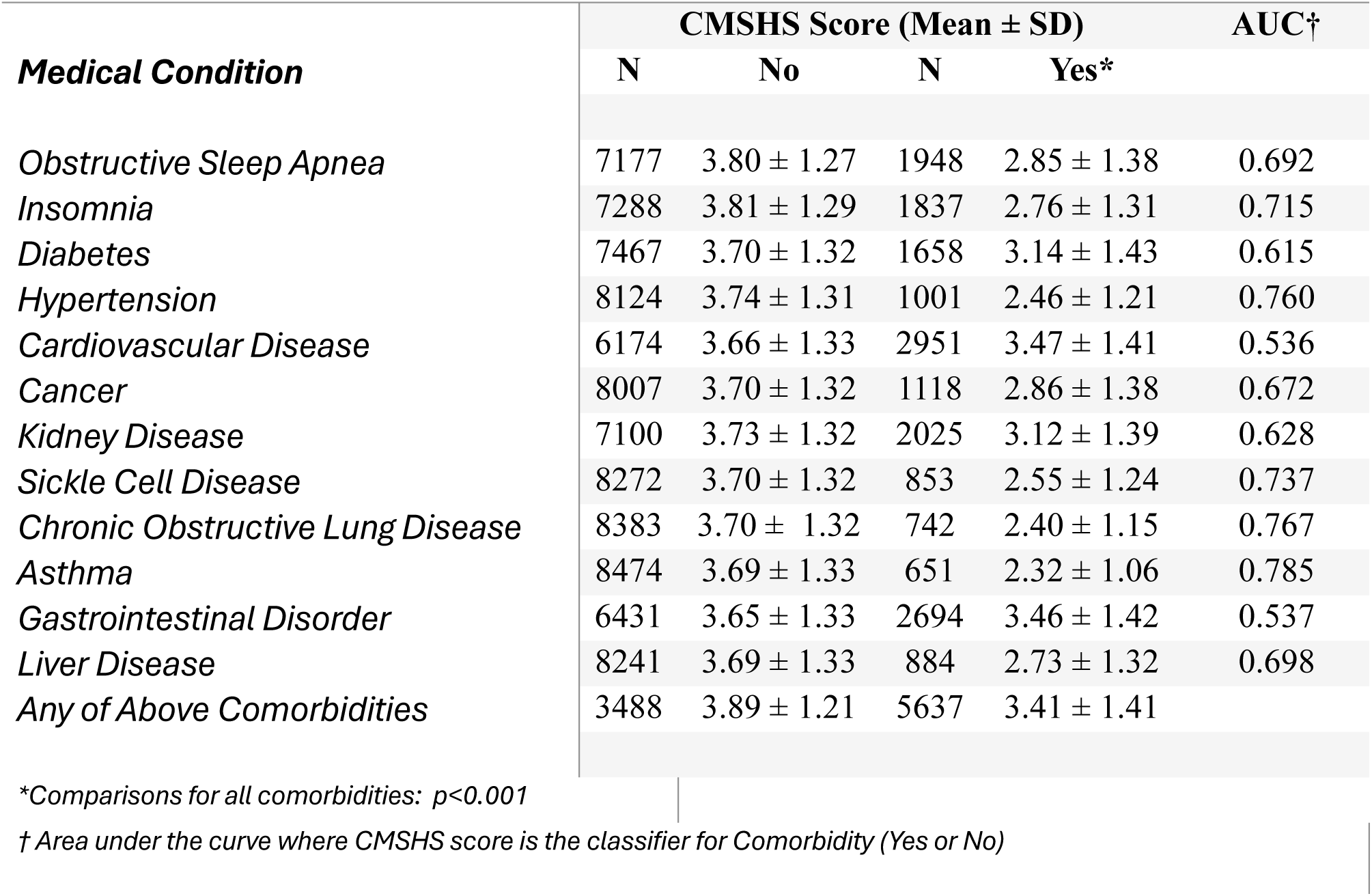
CMSHS Scores for Chronic Medical Conditions.

| Medical Condition | CMSHS Score (Mean ± SD) |  |  |  | AUC† |
| --- | --- | --- | --- | --- | --- |
|  | N | No | N | Yes* |  |
| Obstructive Sleep Apnea | 7177 | 3.80 ± 1.27 | 1948 | 2.85 ± 1.38 | 0.692 |
| Insomnia | 7288 | 3.81 ± 1.29 | 1837 | 2.76 ± 1.31 | 0.715 |
| Diabetes | 7467 | 3.70 ± 1.32 | 1658 | 3.14 ± 1.43 | 0.615 |
| Hypertension | 8124 | 3.74 ± 1.31 | 1001 | 2.46 ± 1.21 | 0.760 |
| Cardiovascular Disease | 6174 | 3.66 ± 1.33 | 2951 | 3.47 ± 1.41 | 0.536 |
| Cancer | 8007 | 3.70 ± 1.32 | 1118 | 2.86 ± 1.38 | 0.672 |
| Kidney Disease | 7100 | 3.73 ± 1.32 | 2025 | 3.12 ± 1.39 | 0.628 |
| Sickle Cell Disease | 8272 | 3.70 ± 1.32 | 853 | 2.55 ± 1.24 | 0.737 |
| Chronic Obstructive Lung Disease | 8383 | 3.70 ± 1.32 | 742 | 2.40 ± 1.15 | 0.767 |
| Asthma | 8474 | 3.69 ± 1.33 | 651 | 2.32 ± 1.06 | 0.785 |
| Gastrointestinal Disorder | 6431 | 3.65 ± 1.33 | 2694 | 3.46 ± 1.42 | 0.537 |
| Liver Disease | 8241 | 3.69 ± 1.33 | 884 | 2.73 ± 1.32 | 0.698 |
| Any of Above Comorbidities | 3488 | 3.89 ± 1.21 | 5637 | 3.41 ± 1.41 |  |

\*Comparisons for all comorbidities: $p < 0.001$
† Area under the curve where CMSHS score is the classifier for Comorbidity (Yes or No)

Comparisons of the CMSHS scores between participants with one COVID-19 infection and those with no history of a COVID-19 infection are shown in Table 3. Participants with a history of COVID-19 had a lower CMSHS score than those who had not been infected (3.52 ± 1.37 vs. 3.78 ± 1.30 p<0.001). Furthermore, infected participants had a smaller proportion of scores ≥ 3 (76.5% vs. 83.1, p<0.001) and ≥ 4 (54.1% vs. 63.2, p<0.001). With the exception of sleep duration, all of the components of the CMSHS score had a smaller proportion of scores in the acceptable range in comparison to those with previous COVID-19.

**Table 3:** Comparison of CMSHS Scores: No COVID-19 Infection vs. One COVID-19 Infection.

|  | Overall* |  | No COVID-19 Infection |  | One COVID-19 Infection |  | p |
| --- | --- | --- | --- | --- | --- | --- | --- |
|  | N=9125 |  | N=5574 |  | N=4328 |  |  |
| CMSHS (Mean ± SD) | 9125 | 3.59 ± 1.36 | 4556 | 3.78 ± 1.30 | 3471 | 3.52 ± 1.37 | <0.001 |
|  | N | % Prevalence | N | % Prevalence | N | % Prevalence |  |
| CMSHS | 9125 |  | 4556 |  | 3471 |  |  |
| 0 | 109 | 1.2 | 51 | 1.1 | 42 | 1.2 |  |
| 1 | 591 | 6.5 | 191 | 4.2 | 623 | 7.6 |  |
| 2 | 1276 | 14.0 | 526 | 11.5 | 511 | 14.7 | <0.001 |
| 3 | 1979 | 21.7 | 908 | 19.9 | 778 | 22.4 |  |
| 4 | 2654 | 29.1 | 1456 | 32.0 | 970 | 27.9 |  |
| 5 | 1983 | 21.7 | 1119 | 24.6 | 714 | 20.6 |  |
| 6 | 533 | 5.8 | 305 | 6.7 | 193 | 5.6 |  |
| CMSHS Cut point $\geq 3$ | 9125 | | 4556 | | 3471 | | |
| <3 | 1976 | 21.7 | 768 | 16.9 | 816 | 23.5 | <0.001 |
| $\geq 3$ | 7149 | 78.3 | 3788 | 83.1 | 2655 | 76.5 | |
| CMSHS Cut point $\geq 4$ | 9125 | | 4556 | | 3471 | | |
| <4 | 3955 | 43.3 | 1656 | 36.8 | 1594 | 45.9 | <0.001 |
| $\geq 4$ | 5170 | 56.7 | 2880 | 63.2 | 1877 | 54.1 | |
| Regularity |  |  |  |  |  |  |  |
| Yes | 7903 | 71.0 | 4242 | 77.6 | 2899 | 68.1 | <0.001 |
| No | 3226 | 29.0 | 1224 | 22.4 | 1355 | 31.9 |  |
| Satisfaction (Quality) |  |  |  |  |  |  |  |
| Yes | 8145 | 71.9 | 4114 | 73.9 | 3046 | 70.2 | <0.001 |
| No | 3178 | 28.1 | 1450 | 26.1 | 1292 | 29.8 |  |
| Alertness |  |  |  |  |  |  |  |
| Yes | 8898 | 80.1 | 4655 | 85.4 | 3305 | 77.8 | <0.001 |
| No | 2209 | 19.9 | 799 | 14.6 | 945 | 22.2 |  |
| Timing |  |  |  |  |  |  |  |
| Yes | 2882 | 25.5 | 1513 | 27.4 | 1073 | 24.9 | 0.006 |
| No | 8349 | 74.3 | 4010 | 72.6 | 3234 | 75.1 |  |
| Efficiency |  |  |  |  |  |  |  |
| Yes | 4482 | 47.8 | 2430 | 52.0 | 1659 | 46.4 | <0.001 |
| No | 4886 | 52.2 | 2247 | 48.0 | 1917 | 53.6 |  |
| Duration |  |  |  |  |  |  |  |
| Yes | 5941 | 52.5 | 2959 | 53.2 | 2316 | 53.4 | 0.839 |
| No | 5382 | 47.5 | 2605 | 46.8 | 2022 | 46.6 |  |

\*All participants with CMSHS Scores

Using two definitions of PASC, Table 4 shows overall and component CMSHS scores for participants with PASC in comparison to those without PASC. The prevalence of PASC was 26.0% with the more expansive NICE definition and 12.9% with the stricter COPE definition. For both definitions, overall CMSHS scores were on average >1 unit lower and the proportion of participants with CMSHS scores < 3 (45.4 vs. 15.5%, p<0.001) and < 4 (73.6 vs. 35.5%, p<0.001) was higher in participants with PASC. Furthermore, for all 6 components of the CMSHS scores, the proportion not in the acceptable range was greater in participants with PASC regardless of definition.

**Table 4:**
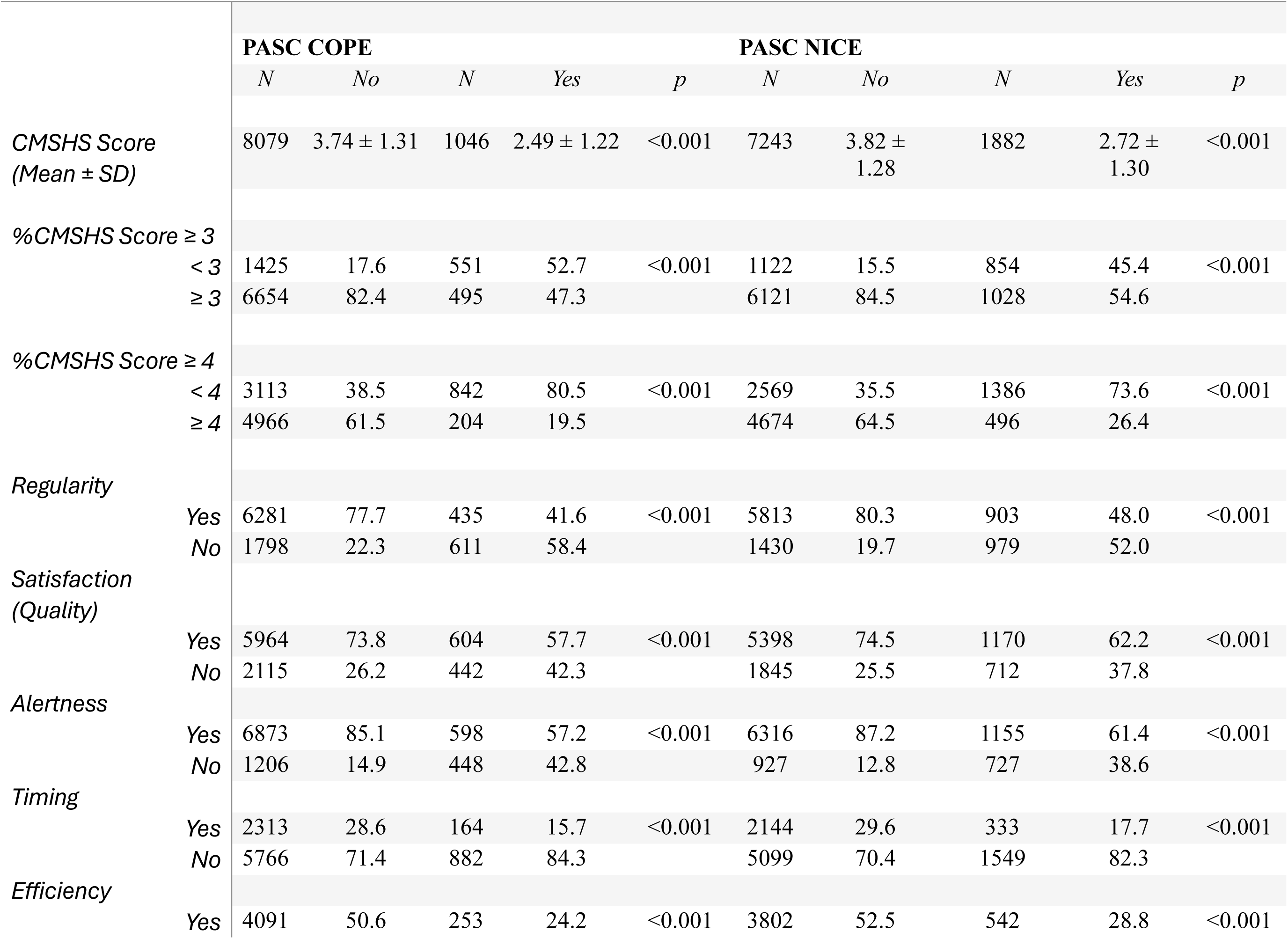
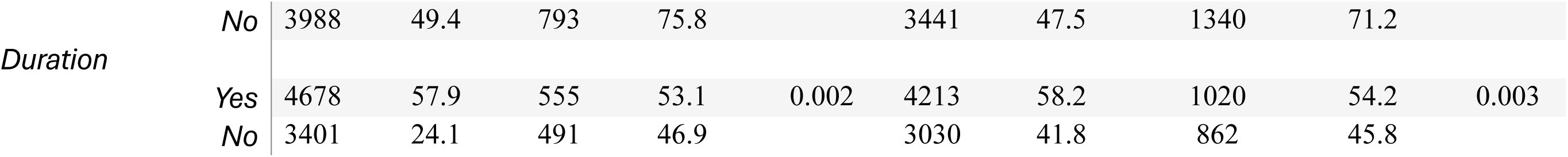
CMSHS Scores in Two Definitions of PASC.

Table 5 shows the changes in CMSHS scores occurring up to 6-12 months after COVID-19 infection in participants who met the criteria for PASC using the both the NICE and COPE definitions. In models unadjusted for covariates, CMSHS scores are lower within 1 month after COVID-19 infection in comparison to participants without PASC (3.49 ± 0.05 vs. 3.86 ± 0.02, p<0.001 for NICE definition; 3.28 ± 0.05 vs. 3.78 ± 0.02, p<0.001 for COPE definition). In those participants who continue to have symptoms, they further decline over the next 3-6 months after which they plateau between 6-12 months (2.66 ± 0.05 vs. 2.60 ± 0.04, p>0.50 for NICE definition; 2.37 ± 0.06 vs. 2.51 ± 0.07, p>0.50 for COPE definition). These associations attenuated after construction of progressively more complex models with covariates, but retained significance for both the NICE definition (3.52 ± 0.05 vs. 2.98 ± 0.04, p<0.001, <1 Month vs. 6-12 Months) and the COPE definition (3.38 ± 0.05 vs. 2.85 ± 0.07, p<0.001, <1 Month vs. 6-12 Months) in fully adjusted models.

**Table 5:**
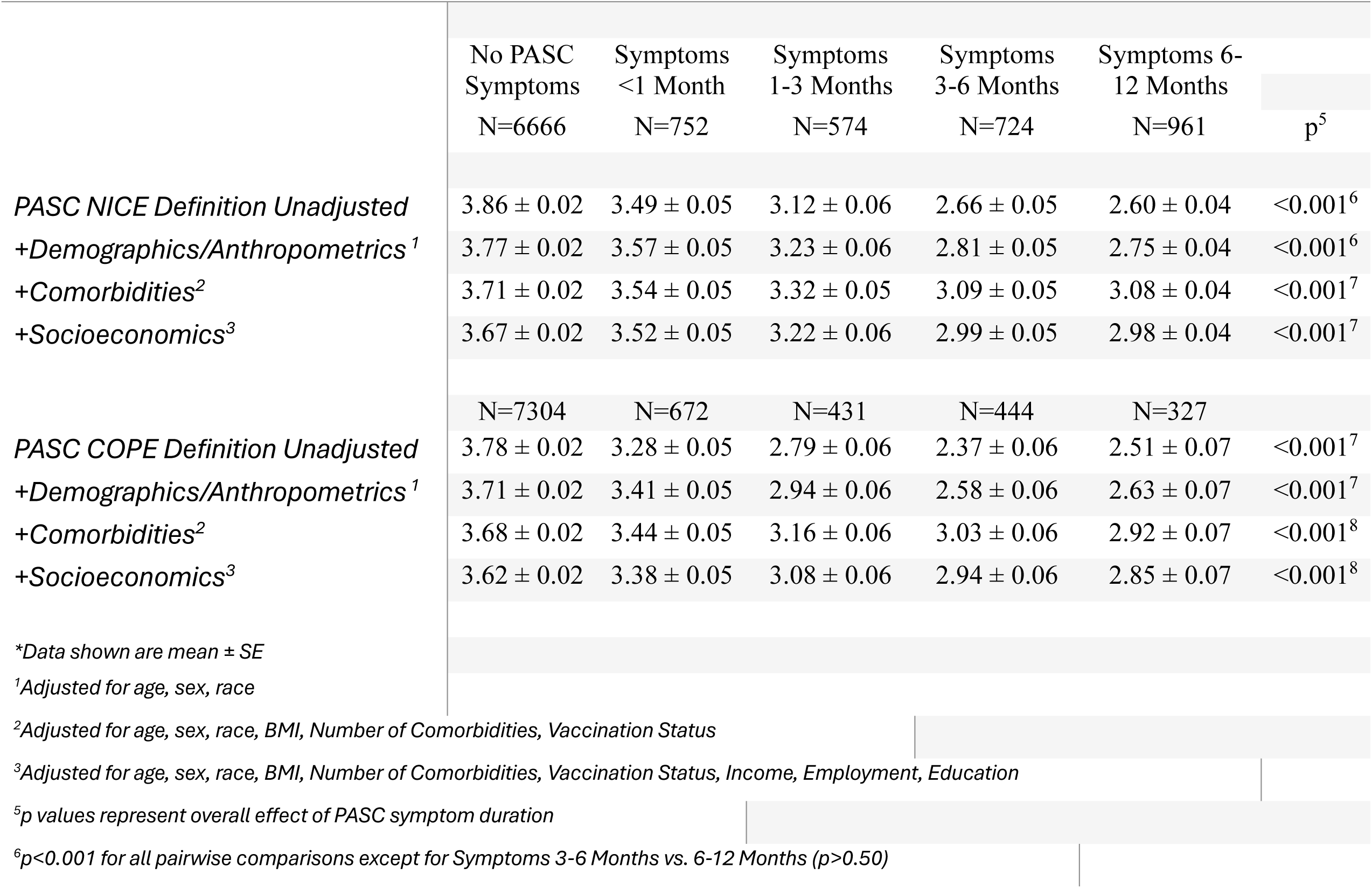
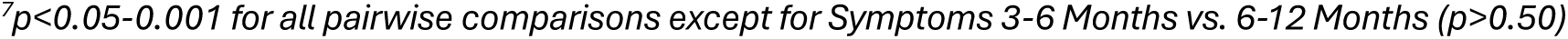
Changes in CMSHS Scores with Increasing Duration of PASC Symptoms*.

|  | No PASC Symptoms<br>N=6666 | Symptoms <1 Month<br>N=752 | Symptoms 1-3 Months<br>N=574 | Symptoms 3-6 Months<br>N=724 | Symptoms 6-12 Months<br>N=961 | p<sup>5</sup> |
| --- | --- | --- | --- | --- | --- | --- |
| *PASC NICE Definition Unadjusted* | 3.86 ± 0.02 | 3.49 ± 0.05 | 3.12 ± 0.06 | 2.66 ± 0.05 | 2.60 ± 0.04 | <0.001<sup>6</sup> |
| *+Demographics/Anthropometrics<sup>1</sup>* | 3.77 ± 0.02 | 3.57 ± 0.05 | 3.23 ± 0.06 | 2.81 ± 0.05 | 2.75 ± 0.04 | <0.001<sup>6</sup> |
| *+Comorbidities<sup>2</sup>* | 3.71 ± 0.02 | 3.54 ± 0.05 | 3.32 ± 0.05 | 3.09 ± 0.05 | 3.08 ± 0.04 | <0.001<sup>7</sup> |
| *+Socioeconomics<sup>3</sup>* | 3.67 ± 0.02 | 3.52 ± 0.05 | 3.22 ± 0.06 | 2.99 ± 0.05 | 2.98 ± 0.04 | <0.001<sup>7</sup> |
|  | N=7304 | N=672 | N=431 | N=444 | N=327 |  |
| *PASC COPE Definition Unadjusted* | 3.78 ± 0.02 | 3.28 ± 0.05 | 2.79 ± 0.06 | 2.37 ± 0.06 | 2.51 ± 0.07 | <0.001<sup>7</sup> |
| *+Demographics/Anthropometrics<sup>1</sup>* | 3.71 ± 0.02 | 3.41 ± 0.05 | 2.94 ± 0.06 | 2.58 ± 0.06 | 2.63 ± 0.07 | <0.001<sup>7</sup> |
| *+Comorbidities<sup>2</sup>* | 3.68 ± 0.02 | 3.44 ± 0.05 | 3.16 ± 0.06 | 3.03 ± 0.06 | 2.92 ± 0.07 | <0.001<sup>8</sup> |
| *+Socioeconomics<sup>3</sup>* | 3.62 ± 0.02 | 3.38 ± 0.05 | 3.08 ± 0.06 | 2.94 ± 0.06 | 2.85 ± 0.07 | <0.001<sup>8</sup> |

\*Data shown are mean ± SE
<sup>1</sup>Adjusted for age, sex, race
<sup>2</sup>Adjusted for age, sex, race, BMI, Number of Comorbidities, Vaccination Status
<sup>3</sup>Adjusted for age, sex, race, BMI, Number of Comorbidities, Vaccination Status, Income, Employment, Education
<sup>5</sup>p values represent overall effect of PASC symptom duration
<sup>6</sup>p<0.001 for all pairwise comparisons except for Symptoms 3-6 Months vs. 6-12 Months (p>0.50)
<sup>7</sup> $p < 0.05$ - $0.001$ for all pairwise comparisons except for Symptoms 3-6 Months vs. 6-12 Months ( $p > 0.50$ )

## Discussion

The current study demonstrates that prior COVID-19 was associated with worse multidimensional sleep health. Using a multidimensional sleep health score with the six domains conveyed by the validated RuSATED scale, we found that persons with prior COVID-19 had lower sleep health scores than those uninfected. Furthermore, a greater adverse effect on sleep health was observed in individuals who met the definition of PASC. In those with PASC, sleep health declined over an initial 6-month period and eventually plateaued over the next 6 months.

We obtained an assessment of overall sleep health using an adaptation of the dichotomous version of the RuSATED questionnaire validated by Brindle et al [12]. The primary modification that we made was to substitute questionnaire data for sleep timing and duration obtained by actigraphy. Although using questionnaire data rather than actigraphy is likely less accurate and has greater variability, good external validation was observed when applied against several chronic medical conditions known to negatively impact sleep health. This suggests that our modification maybe useful in future applications where actigraphy is not available.

Symptoms of sleep disturbances are common in association with COVID-19 infection [5, 6, 8, 19, 20]. In a systematic review, a higher pooled prevalence rate of sleep problems has been reported globally among patients affected by COVID-19 compared to the general population (74.8%; 95% CI, 28.7–95.6% vs 35.7%; 95% CI, 29.4–42.4%).[20] Similarly, another meta-analysis demonstrated an increased pooled prevalence among patients with COVID-19 (57%; 95% CI: 42-72%).[5] A systematic review of literature on Middle Eastern and North African (MENA) populations also demonstrated poor sleep health in association with the COVID-19 pandemic [21]. However, the studies summarized in these meta-analyses and reviews refer to a heterogeneous collection of either specific sleep concerns or dimensions, e.g., sleep duration, or vague terminology, e.g., sleep problem or disturbance. Recently using the SATED and Ru-SATED questionnaires, a repeated cross-sectional study, examined sleep health trends following COVID-19 pandemic and found heterogeneous improvements across sleep health dimensions. The study found improvement in sleep efficiency and timing, whereas sleep satisfaction remained low. However, due to the nature of this study individual level trajectory was not assessed and within-person changes over time could not be evaluated. Additionally, the information regarding COVID-19 infection was not examined and effect of viral infection on sleep health could not be evaluated [14].

To our knowledge, our study is the first to demonstrate that overall multidimensional sleep health is negatively associated with COVID-19 infection. Our finding that five of the six dimensions of sleep health were negatively affected provides evidence that COVID-19 has an adverse impact on almost all aspects of sleep health.

We observed that sleep health was markedly worse in persons who met the definition of PASC. Our finding is consistent with numerous reports of a high prevalence of sleep disturbances in PASC. Although the prevalence of sleep disturbances in PASC varies widely, a meta-analysis of 29 studies reported a global prevalence of 46% (95% CI: 38-54%) [22]. In another systematic review and meta-analysis of 194 prospective studies, sleep disturbance without specificity was noted among 13.78%–29.42% of persons with PASC [3]. However, most studies in this review included hospitalized patients which may have overestimated the prevalence among healthy controls. In a recent report from the RECOVER cohort, 12% reported sleep disturbance (adjusted OR of 3.51 (2.46, 5.02) compared to 4% among noninfected participants [23]. With respect to specific sleep conditions or disorders, insomnia is the most common specific sleep condition reported among participants with PASC. In a recent meta-analysis, the pooled prevalence of insomnia in persons with PASC was 38% [22]. In an online survey of people with COVID-19, 78.6% (95% CI 84.0% to 79.9%) of respondents experienced sleep disturbances; insomnia was reported by 60% in comparison to sleep apnea which was reported by 10% [19]. However, among those with insomnia, 21% had experienced insomnia prior to COVID-19 infection, and 34% of those reporting sleep apnea had preexisting sleep apnea. There also is data from several large population based studies demonstrating that obstructive sleep apnea is a risk factor for PASC [16, 24]. Although by implication sleep health is diminished in these studies, its multidimensional construct was not explored. Thus, we extend the findings of these studies by demonstrating that not only is overall sleep health is reduced in PASC, but that virtually all dimensions of sleep health are negatively impacted as well.

In our analysis, we observed that not only overall sleep health was reduced in persons with PASC, but that it appeared to decline for several months post infection, eventually plateauing after 6-12 months. Other cross-sectional studies have demonstrated persistence of sleep disturbance or insomnia for extended durations after COVID-19 infection [4, 19],[25], but none have assessed multidimensional sleep health. Furthermore, our data are consistent with a recent prospective study from the RECOVER study reporting that persistence of symptoms is one of the eight relatively distinct longitudinal profiles observed with PASC [26], However, the trajectories of neither individual sleep symptoms or conditions, nor overall sleep health were described. Thus, we broaden the findings from the RECOVER study [26] and others [4, 19],[25] by demonstrating that impairments in sleep health may be persistent in some individuals after COVID-19 infection. It is incumbent on clinicians to be aware of this finding and provide supportive care even though definitive treatment remains elusive. Our findings may also guide community-based interventions or public health campaigns to promote awareness about healthy sleep strategies for persons with PASC. Given the role of sleep as one of the 3 pillars of good health [27], further research is needed to identify the optimal therapeutic targets and modalities (e.g., in-person with a healthcare provider, social media, public health campaigns, or other avenues) for improving sleep health in persons with PASC.

Although the exact etiology of the COVID-19 impact on sleep health is not established, several mechanisms have been proposed. Structural changes in the brain have been reported after COVID-19 infection [28]. Thus, there may be direct damage of virus to the central nervous system leading to neuronal death or dysfunction and alterations in neurotransmitters. In addition, COVID-19 infection can lead to persistent inflammatory responses, immune activation, mitochondrial dysfunction, and dysregulation of the autonomic nervous system [29–32]. These consequences of infection have been associated with sleep disturbances. The constellation of COVID-19, fever, cough, and difficulty breathing, also contributed to poor sleep. In addition, social isolation, change in sleep-wake schedule, financial impact, and in particular, anxiety related to the fear of infection may have contributed to worse sleep [33],[34], In response to an online survey in March 2020 after the COVID 19 outbreak more than 60% of participants reported anxiety and over half of them also reported sleep disturbance [35]. Similarly, researchers in China observed that during the pandemic individuals were more likely to report insomnia and this also was attributed to anxiety [36].

Despite its strengths, we acknowledge some limitations of this study. Inasmuch as the study is based on self-reported sleep measures, we cannot exclude reporting bias. Additionally, self-report measures lack precision that may have resulted in greater variability in our findings. Future studies using objective sleep measures are needed to better assess sleep health among persons with PASC. Finally, our analyses were cross-sectional, and causality cannot necessarily be inferred.

In conclusion, multidimensional sleep health is negatively associated with COVID-19 infection; individuals with PASC have a greater reduction sleep health that may be persistent for up to 12 months post infection. Additional research to identify effective treatment to improve sleep health after COVID-19 infection is indicated.

## Data Availability

All data produced in the present study are available upon reasonable request to the authors

## Statements and Declarations

### Ethics Approval

All procedures were in accordance with the ethical standards of Monash University Human Research Ethics Committee (Study #24036) and with the 1964 Helsinki declaration and its later amendments or comparable ethical standards. Informed consent was obtained electronically from all individual participants included in the study.

### Author Approval and Consent to Publish

All authors have seen and approved the manuscript; all authors have given their consent to publish.

---

**Table S1:**
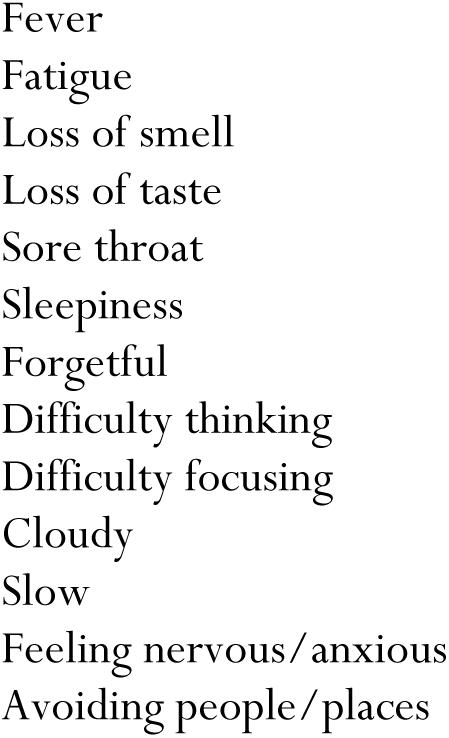
Symptoms Associated with PASC.

